# Equipment Downtime and the Absence of Standardized Practices in Ghanaian Prosthetics and Orthotics Clinics: A Narrative Report from Field Observations

**DOI:** 10.1101/2025.08.17.25333853

**Authors:** Daniel Opoku-Gyamfi, Akouetevi Aduayom-Ahego, Christian Kwaku Agbewoavi, Cecil Owusu Bempah, Joseph Pitoor Bikala, Deborah Adiki Aniteye, Prince Oduro, Harriet Amakye Ansah, Michael Kuaku Bansah, Zenith Yamoah

**Affiliations:** Department of Prosthetics and Orthotics, School of Allied Health Sciences, University of Health and Allied Sciences, Ho, Ghana; New World Company, Osaka, Japan; Department of Prosthetics and Orthotics, Komfo Anokye Teaching Hospital, Kumasi, Ghana; National of Prosthetics and Orthotics Center, Ghana Health Service, Accra, Ghana; Regional Prosthetics and Orthotics Center, Ghana Health Service, Ho, Ghana; Department of Prosthetics and Orthotics, St. Joseph Hospital, Koforidua, Ghana; Department of Physiotherapy, Ho Teaching Hospital, Ho, Ghana; Kekeli Foundation, Ho, Ghana

**Keywords:** Equipment Downtime, Standard Operating Procedures (SOPs), Prosthetics and Orthotics (P&O), Maintenance Gaps, Ghana

## Abstract

**Background:** Prosthetics and Orthotics (P&O) services are essential for restoring mobility and independence, yet in Ghana, access remains limited and inequitable. While recent advances in training and regulation have been achieved, systemic gaps in infrastructure, equipment maintenance, and standardization persist. This study explores the operational realities within Ghana’s P&O clinics to uncover the scale and impact of these challenges.

**Methods:** A qualitative descriptive field observation design was employed across all 14 operational P&O facilities in Ghana from January to April 2025. A multidisciplinary team of licensed professionals conducted week-long visits, using semi-structured observation tools to assess equipment functionality, the presence of Standard Operating Procedures (SOPs), and impacts on patient care and training. Data were collected through real-time handwritten notes and informal discussions, then thematically analyzed.

**Results:** Twelve of the 14 facilities experienced prolonged equipment downtime, with critical machines like vacuum pumps, ovens, and routers non-functional for months or years due to lack of spare parts, local technical expertise, and maintenance planning. No facility used modern technologies such as CAD/CAM. Additionally, no clinic had documented SOPs for clinical assessment, device fabrication, infection control, or follow-up care. This absence of standardization led to inconsistent service quality, compromised patient outcomes, and hindered effective student training.

**Conclusion:** Ghana’s P&O sector is constrained by systemic failures in equipment maintenance and procedural standardization, rooted in underfunding, lack of policy, and technical capacity. These findings mirror challenges across low-resource settings. Addressing these issues through national SOPs, local technician training, and sustainable maintenance systems is essential to improve service delivery, patient outcomes, and the professional readiness of future P&O practitioners.

## Introduction

Prosthetics and Orthotics (P&O) services are an essential component of rehabilitation, enabling individuals with limb loss or musculoskeletal impairments to regain mobility, independence, and quality of life [1]. In low- and middle-income countries (LMICs), demand for these services is rising due to an increasing burden of trauma, diabetes-related amputations, and congenital conditions [2]. Despite this need, global estimates suggest that only 5–15% of people who require assistive devices in LMICs have access to them [3].

In Ghana, the P&O sector has seen modest but important progress in the last decade, with advances in professional training and regulatory oversight. The establishment of the Brother Tarcisius Prosthetics and Orthotics Training College in 2013 and the launch of the first Bachelor’s degree program in P&O at the University of Health and Allied Sciences in 2020 represent critical milestones [4]. Service expansion has been geographically uneven, concentrating facilities in a few regions and widening urban–rural access gaps [5].

Existing literature on P&O services in Ghana has primarily focused on education, workforce development, and broad service capacity [4,6,7]. Very little has been done about the operational realities within clinics, particularly regarding equipment downtime, maintenance practices, and the practices of standardized operational protocols These factors are critical because P&O services are heavily dependent on specialized machinery, such as vacuum systems, alignment jigs, and grinders, that are often imported, expensive to maintain, and difficult to repair locally [6,8]. The failure of such equipment can halt service provision entirely, delay patient care, and undermine both clinical outcomes and staff morale [9,10].

Moreover, while the WHO and the International Society for Prosthetics and Orthotics (ISPO) have developed global standards for P&O service provision, including guidelines on equipment maintenance and quality management, there has been no documented assessment of how (or if) these standards are implemented in Ghana [11,12]. The lack of such data creates a blind spot in both national policy planning and international support initiatives.

This narrative report addresses this gap by providing first-hand field observations from within the Ghanaian P&O service landscape. To our knowledge, this is the first account to document the scale and impact of equipment downtime and the systemic absence of standardized maintenance and operational practices across multiple P&O clinics in Ghana. This report adds a layer of contextual detail absent from previous surveys or policy reviews by grounding the analysis in real-world, on-site experiences. The findings are intended to inform national stakeholders, development partners, and professional bodies seeking to strengthen the resilience, efficiency, and equity of P&O services in Ghana.

## Methodology

### Study design

Qualitative descriptive field observation design was employed.

### Study Setting and Sampling

The study included all 14 operational prosthetics and orthotics facilities in Ghana during the data collection period (January to April 2025), comprising 11 publicly funded and 3 privately operated facilities. Facilities were selected based on their active provision of clinical P&O services and official recognition by the Ministry of Health, Ghana Health Service, and the Allied Health Professions Council, Ghana. Accessibility to the research team during the study period was a key inclusion criterion. Facilities were excluded if they were non-operational, under construction, temporarily closed, or provided only general rehabilitation services without dedicated P&O components. Mobile or outreach services lacking fixed clinic infrastructure were also excluded to maintain focus on established P&O clinical environments.

### Research Team and Training

Data collection was conducted by a multidisciplinary team of eight licensed prosthetists and orthotists with extensive knowledge of Ghana’s P&O sector. To ensure consistency, the team underwent a one-day training session prior to data collection, focusing on the observational framework, ethical considerations, and standardized note-taking procedures. The team was divided into two groups of four members each, with each group assigned to visit seven facilities to balance workload and ensure thorough data collection.

### Data Collection Procedures

Data collection occurred between January and April 2025, with each of the 14 facilities receiving a one-week site visit. Observations were conducted during standard working hours (9:00 AM to 3:00 PM daily) to capture routine clinical operations. Written approval was obtained from facility heads prior to each visit, ensuring cooperation and minimizing disruptions to clinical activities.

A semi-structured observational framework guided data collection, focusing on three thematic areas: (1) Equipment Functionality and Downtime: Assessing the availability, condition, and maintenance of key technical equipment (e.g., grinders, routers, ovens, vacuum systems). (2) Presence of Standard Operating Procedures (SOPs): Evaluating the existence and application of documented clinical and technical protocols for patient assessment, device fabrication, and service delivery. (3) Impact on Patient Care and Clinical Training: Examining how infrastructural gaps and procedural inconsistencies affect service quality and the effectiveness of educational mandates in facilities involved in student training.

Observations were recorded in real-time using handwritten field notes in standardized templates to ensure consistency. Informal discussions with clinic personnel were conducted as needed to clarify observations, with responses documented anonymously to protect confidentiality. To minimize disruption and uphold ethical standards, no photographs or audio recordings were taken. All field notes were transcribed into digital formats using Microsoft Word and Excel within 24 hours of each visit to ensure accuracy and fidelity. Each facility was assigned an anonymized identifier (Facility A to Facility N) to maintain confidentiality throughout data processing and reporting.

### Data Analysis

Qualitative data were analyzed using thematic content analysis, following the six-phase approach outlined by Braun and Clarke (2021) [13]. First, the research team familiarized themselves with the data through independent review of transcribed field notes. Second, initial codes were manually assigned to data segments to identify recurring patterns related to equipment availability, functionality, maintenance practices, clinical workflows, and SOP adherence. Third, codes were grouped into broader thematic categories through iterative discussions. Each team held separate meetings to refine themes, with a designated third researcher per team mediating any coding disagreements to ensure consensus. Fourth, the two mediators collaborated to harmonize themes across teams, ensuring interpretive consistency. Fifth, themes were defined and named to reflect systemic gaps and their implications. Sixth, findings were contextualized within the P&O sector’s operational landscape. Comparative analysis across all facilities identified variations, similarities, and outliers in practices. Quantitative summaries (e.g., number of facilities with equipment breakdowns or lacking SOPs) were used descriptively to complement qualitative findings, but the primary focus remained on rich, contextualized insights into systemic gaps and their implications for P&O service delivery and training.

To enhance credibility, preliminary findings were shared with three facility heads for respondent validation, and no contradictory feedback was received. Nightly debriefing sessions among observers were held to reflect on observations, minimize individual bias, and ensure alignment with the study’s objectives. No specialized software was used for coding; analysis relied on the team’s domain expertise in P&O practice.

### Ethical Considerations

Ethical approval was obtained from the University of Health and Allied Sciences Research Ethics Committee (UHAS-REC, protocol ID UHAS-REC.123.33). Stringent measures ensured data security and confidentiality. No personally identifiable information (PII) from staff or patients was collected. Digital files were stored in password-protected folders on secure local drives, accessible only to the core research team, with encrypted backups maintained on separate storage devices. Physical field notes were stored in locked cabinets when not in use. The study adhered to qualitative descriptive principles, prioritizing thematic synthesis over statistical analysis to highlight systemic issues in Ghana’s P&O sector.

### Trustworthiness

To ensure trustworthiness, the study employed multiple strategies. Credibility was enhanced through prolonged engagement (one-week visits per facility), respondent validation, and triangulation of data from observations and informal discussions. Dependability was supported by detailed documentation of the research process, including the observational framework and coding procedures. Confirmability was achieved through nightly debriefs and mediator involvement to minimize bias. Transferability was facilitated by thick descriptions of the study context and anonymized facility profiles, enabling readers to assess the applicability of findings to similar settings.

## Results

As part of our assessment of prosthetics and orthotics (P&O) service delivery in Ghana, we visited all 14 operational facilities across the country as of 31st July 2025. These visits were aimed at gathering firsthand insights into the operational realities on the ground. The observations documented in this section reflect a wide range of challenges and gaps, particularly related to equipment availability and maintenance, as well as the lack of standardized practices across facilities. To preserve anonymity while still conveying critical details, the clinics observed are referred to as Facility A, Facility B, and so on. The findings are organized into two main themes that emerged consistently during the visits: Equipment Downtime and Maintenance Gaps, and Absence of Standard Operating Procedures (SOPs).

### Current state of Prosthetics and Orthotics service in Ghana

Prosthetics and Orthotics (P&O) services in Ghana are delivered through a mix of public and private institutions, with varying capacities and infrastructure. The P&O sector, although essential in providing mobility solutions and rehabilitation care, remains relatively underdeveloped compared to other healthcare fields in the country. The general workflow within these clinics includes patient assessment, device fabrication, fitting, and follow-up services relying heavily on specialized equipment and skilled professionals.

As of 31st July 2025, Ghana had a total of 14 functional P&O facilities. These facilities are not equitably distributed across the country. Out of the 16 administrative regions in Ghana, only 7 regions currently host P&O centers. A significant concentration (n=12) is found in the southern and central parts of the country, namely Greater Accra (4), Eastern (3), Central (2), Ashanti (2), and Ahafo (1). In contrast, only two facilities are located in the northern regions—Upper East and North East, each hosting just one center.

Importantly, there is currently only one active P&O clinic within one of Ghana’s teaching hospitals. Also, only one P&O clinic exist in one of the regional hospitals in Ghana. The majority of the P&O facilities in Ghana (n= 9) are standalone clinics, often not integrated into broader hospital systems, which may limit interdisciplinary care and access to emergency support. Among the 14 facilities, 3 are privately owned, with the rest under various government or mission-based management.

In terms of training and professional development, the Brother Tarcisius Prosthetics and Orthotics Training College in Nsawam, Eastern Region, was the first institution in Ghana to offer formal education in the field, launching its Diploma and Certificate programs in September 2013. A significant milestone was achieved in 2020, when the University of Health and Allied Sciences (UHAS) began the first Bachelor’s degree program in Prosthetics and Orthotics in Ghana. The inaugural cohort graduated in November 2024, marking a key development in local professional training. Before this, Ghanaians with degree-level training in P&O had obtained their qualifications outside the country or through upgrading programs facilitated by international collaborations, such as those run by Human Study e.V.

In Ghana, all P&O professionals are licensed by the Allied Health Professions Council (AHPC). To be eligible for licensure, graduates must first complete a one-year mandatory internship, which also serves as their national service to the country. Upon successful completion, candidates sit for the licensing examination administered by the AHPC. As of July 2025, the pass mark for the licensing exam is 60%.

Despite these advancements in education and regulation, standardized procedures for equipment maintenance, service quality, and facility management are still lacking nationwide. These gaps have real implications for the effectiveness and sustainability of care provided to individuals who rely on prosthetic and orthotic devices.

In a typical P&O clinic, the workflow involves patient assessment, casting, fabrication, alignment, and fitting, followed by gait training and follow-up. Each step is heavily reliant on a range of equipment; from vacuum suction machines, alignment jigs, and grinders to ovens, routers, and stitching machines. Most of these tools are imported, expensive to maintain, and often lack local servicing options. When a key machine breaks down, there is usually no backup. Clinics are forced to improvise, refer patients elsewhere, or pause services entirely. As of July 2025, no facility actively use new technologies like CAD/CAM for clinical services. All facilities still engage in traditional method of practice.

While there have been occasional efforts, mainly donor-driven or ad hoc internal initiatives, to standardize clinical practices or improve infrastructure, these have not been sustained or scaled nationally. There is currently no comprehensive national policy or maintenance framework guiding the upkeep and standardization of equipment across clinics. As a result, service delivery varies significantly from one facility to another, affecting patient outcomes and professional morale alike.

### Equipment Downtime and Maintenance Gaps

Across the spectrum of P&O clinics, from larger urban centers to smaller regional facilities, a common array of specialized machinery forms the backbone of fabrication workshops. All facilities use basic equipment such as grinders, routers, vacuum systems, ovens for thermoplastic forming, laminating stations, drill presses, and various hand tools. These machines are indispensable for the precise shaping, molding, and finishing of custom prosthetic limbs and orthotic braces, directly influencing the fit, comfort, and functionality of the devices provided to patients.

Despite their fundamental importance, a striking and concerning pattern of non-functional equipment emerged during the field observations. At least 12 of the 14 facilities had one or more major equipment non-functional at the time of visit. This was not merely a matter of minor malfunctions but often involved critical pieces of machinery that were essential for core fabrication processes. The sight of idle, often dust-covered, and disconnected equipment was a recurring feature in workshops, serving as a constant reminder of the operational limitations faced by these clinics.

The severity of this issue was further underscored by the duration of downtime observed. It was not uncommon for several machines to have been down for months or even years, with no clear maintenance plan in place. For instance, a heavy-duty router at facility A had been inoperable for nearly eighteen months, with its cutting head visibly corroded and its power supply disconnected. Similarly, a crucial vacuum pump at the same facility A, vital for both thermoplastic molding and lamination, had been out of commission for over a year, forcing technicians to resort to laborious and less precise manual methods. This prolonged idleness of essential equipment pointed to a systemic inability to address breakdowns in a timely or effective manner. Also, facility C’s large, industrial-grade thermovacuum forming oven, a cornerstone for fabricating custom thermoplastic orthoses, was found to be completely non-functional. According to staff, this critical piece of equipment had been out of commission for over two years. The primary reason cited was the unavailability of a specific heating element and control board, coupled with the absence of local technicians possessing the specialized knowledge required for its repair. Attempts to source spare parts internationally were reportedly hampered by prohibitive costs and complex import logistics. Consequently, the facility was forced to resort to less efficient and less precise alternative methods, such as manual heating with heat guns or outsourcing certain fabrication steps, which significantly increased production time and potentially compromised the consistency and quality of the final devices.

Similarly, at Facility D, a crucial vacuum pump, essential for both thermoplastic forming and lamination, had ceased to function approximately eight months prior to the observation. This particular pump was noted to be an older model, making the procurement of compatible spare parts exceedingly difficult. Without a functional vacuum pump, the clinic’s ability to create custom-fitted sockets was severely curtailed. Technicians were observed attempting to achieve some level of vacuum using improvised methods or relying on manual pressure, which, while demonstrating remarkable ingenuity, could not replicate the consistent and precise suction required for optimal material conformity. This often resulted in less intimate fits for patients, requiring more adjustments and potentially leading to discomfort or reduced functionality of the prosthetic or orthotic device.

Beyond these specific examples, general observations across other unnamed facilities underscored a systemic issue. Grinding machines frequently operated with worn-out or incorrect abrasive wheels, if they operated at all, leading to inefficient material removal and a rougher finish on devices. Drill presses often lacked proper bits or were unstable, posing safety concerns and limiting precision. The common threads linking these instances of equipment downtime were a critical shortage of readily available spare parts, a severe dearth of locally trained maintenance technicians capable of handling specialized P&O machinery, and often, insufficient budgetary allocations for proactive maintenance or timely repairs. The reliance on foreign technicians for complex repairs meant lengthy delays and exorbitant costs, often making repair an unfeasible option for underfunded clinics.

The underlying causes for this widespread equipment failure were consistently identified across the visited sites. The most prominent factors included a pervasive lack of spare parts, often exacerbated by the fact that much of the equipment is imported, making sourcing specific components difficult and expensive. This was coupled with a critical absence of local technical support; there were very few, if any, trained biomedical engineers or specialized technicians within Ghana capable of diagnosing and repairing complex P&O machinery. Clinics often reported having to wait indefinitely for foreign technicians or attempting improvised, often unsuccessful, repairs themselves. Furthermore, a general lack of understanding or implementation of poor preventive maintenance practices contributed significantly to premature equipment failure. Routine cleaning, lubrication, and calibration, which could extend the lifespan of these machines, were rarely observed or systematically performed.

To illustrate the tangible impact of these issues, Facility G, a medium-sized clinic serving a significant patient population. During the visit, the facility’s primary vacuum pump, a cornerstone for creating well-fitting prosthetic sockets, had been out of use for over 2 years. This forced the technicians to resort to entirely manual fabrication of sockets. Instead of using the controlled environment of a vacuum former to draw thermoplastic sheets tightly over patient molds, they were observed manually pressing the heated plastic, attempting to achieve an adequate fit. This process was not only physically demanding and time-consuming but also introduced significant variability in socket quality, often resulting in less intimate fits, increased pressure points, and a higher likelihood of needing multiple adjustments or even complete refabrication. The absence of this single piece of equipment severely compromised the precision and efficiency of their entire socket production line. In another instance at Facility K, the main buffing machine, essential for smoothing and finishing devices, had a broken motor for over six months. This meant that all finishing work had to be done laboriously by hand, extending production times and often resulting in a less refined aesthetic finish on the final devices.

The ramifications of this widespread equipment downtime are multifaceted and deeply detrimental. For patient care, the most immediate effect is a significant increase in waiting times. Fabrication processes that should take days can stretch into weeks or even months due to the need for manual workarounds or the inability to perform certain steps. This delay can have profound impacts on patients’ rehabilitation journeys, prolonging their period of immobility or discomfort. Furthermore, the inability to utilize proper machinery often leads to compromises in the quality, fit, and aesthetic finish of the prosthetic and orthotic devices. Patients may receive devices that are less comfortable, less durable, or less functionally appropriate than what could be achieved with fully operational equipment, potentially leading to dissatisfaction, abandonment of devices, and poorer long-term outcomes.

Equally concerning is the impact on student training. Ghanaian P&O training programs aim to produce competent professionals capable of operating modern equipment. However, when students are trained in environments where essential machines are perpetually broken, their practical exposure is severely limited. They graduate with theoretical knowledge but often lack hands-on proficiency in using the very tools that are standard in more developed P&O settings. This creates a significant skill gap, making it challenging for them to integrate effectively into well-equipped clinics, or to establish high-standard practices themselves. Moreover, witnessing the constant struggle with broken equipment can be profoundly demotivating for aspiring P&O professionals, potentially leading to a sense of disillusionment about the feasibility of providing optimal care within the existing infrastructure. The cycle perpetuates itself, as a lack of skilled technicians to maintain equipment contributes to the very problem that hinders the training of future technicians.

### Absence of Standard Operating Procedures (SOPs)

Beyond the tangible challenges posed by equipment downtime, a more subtle yet equally pervasive issue observed across all the 14 Ghanaian P&O clinics was the conspicuous absence of formalized Standard Operating Procedures (SOPs). This lack of standardized protocols extended from the initial patient assessment and casting stages through to fabrication, fitting, follow-up care and workshop safety procedures. The prevailing approach appeared to be one of individual discretion and reliance on ad-hoc methods, largely shaped by the training background and personal preferences of the lead prosthetist or orthotist at each facility. There was no evidence of nationally recognized protocols, comprehensive technical manuals, or universally adopted best practices guiding the day-to-day operations. The following are the identified common areas lacking SOPs across the 14 P&O clinics.

#### Clinical Assessment and Documentation

A critical foundational step in P&O clinical practice, patient evaluation, exhibited remarkable variability across all visited sites. None of the facilities had documented protocols guiding patient evaluation, diagnostic documentation, or clinical decision-making pathways. The process was overwhelmingly reliant on the individual clinician’s experience, training background, and personal judgment. For instance, the specific anatomical landmarks used for measurements, the method of recording patient history, or the systematic assessment of functional limitations varied significantly across all the 14 P&O clinics. Patient files, where they existed (n=6), often lacked uniformity in their structure and content, making it challenging to track comprehensive patient journeys, monitor progress, or ensure continuity of care, especially if a patient moved between facilities or was attended by different clinicians. Clinical decisions, such as the choice of a specific device type or material, were often made without reference to a standardized decision-making tree or evidence-based guidelines, instead stemming from the practitioner’s accumulated practical knowledge.

#### Device Fabrication and Technical Processes

The fabrication of custom P&O devices, was another area where standardization was conspicuously absent. There were no standard procedures for measurements, alignment, casting, or component selection, which led to observed variability in technical practice. For example, the technique for taking a plaster cast of a residual limb, including the specific plaster-to-water ratio and drying time differed from one technician to another. Similarly, the methods for modifying the positive mold (e.g., pressure relief areas, build-ups) were subjective, based on individual interpretation rather than a universal standard. The critical process of prosthetic alignment, which dictates gait efficiency and patient comfort, lacked any documented static or dynamic alignment protocols. Technicians often relied on visual cues and iterative adjustments without a systematic, easy-reproducible method. Even the selection of prosthetic components (e.g., feet, knees, ankle joints) was often driven by availability or individual preference rather than a standardized assessment of patient needs, activity levels, and biomechanical requirements. This lack of uniformity directly translated into observable differences in the quality, fit, and functional performance of devices produced across, and sometimes even within, different facilities.

Concerns regarding safety were also amplified by the lack of standardization. Without clear guidelines for material selection, fabrication techniques, and quality control checks, there was an inherent risk of producing devices that might not meet essential safety standards. For example, the curing process for resins used in lamination, which dictates the strength and stability of the final product, was not consistently monitored or documented. Variations in temperature, humidity, or mixing ratios, if not controlled by an SOP, could lead to compromised material properties, increasing the risk of device failure during use. Similarly, the absence of standardized protocols for strap attachment, padding application, and component assembly meant that critical safety features might be overlooked or inconsistently applied, potentially leading to skin breakdown, falls, or other injuries for the patient. A specific instance highlighting this issue involved the fabrication of orthotic ankle-foot orthoses (AFOs). In one clinic, the AFOs were consistently observed to have well-defined trim lines and smooth edges, indicating careful finishing. However, in another facility, AFOs occasionally presented with rougher edges or inconsistent trim lines, necessitating further adjustments during fitting or potentially causing discomfort for the patient. This discrepancy was not due to a lack of skill but rather the absence of a shared, documented methodology for achieving a consistent, high-quality finish. The reliance on individual experience, while valuable, could not substitute for the systematic consistency that robust SOPs provide, particularly in ensuring uniform quality and safety across all fabricated devices.

Furthermore, the process of alignment, the precise positioning of prosthetic components to ensure optimal gait and balance, was often highly subjective. While experienced prosthetists possess an intuitive understanding of biomechanics, the absence of standardized static and dynamic alignment protocols meant that the final alignment of a prosthesis was heavily dependent on the individual’s judgment rather than a reproducible, evidence-based process. This led to observable inconsistencies in patient gait patterns. Some patients exhibited noticeable deviations, such as excessive trunk lean or circumduction, which could be attributed to suboptimal alignment that might have been mitigated by adherence to a standardized protocol. This not only affected the patient’s mobility but also placed undue stress on other joints and muscles, potentially leading to secondary musculoskeletal issues over time.

#### Infection Prevention and Workshop Safety

One of the most concerning gaps was the absence of formalized procedures for maintaining a safe and hygienic environment. No facilities demonstrated written SOPs for infection control, waste disposal, or occupational safety measures within the clinical or technical spaces. While some general awareness of hygiene was present, there were no clear guidelines for sterilizing tools, managing clinical waste (e.g., bandages, stockings), or handling hazardous materials used in fabrication (e.g., resins, solvents). Workshops, which involve power tools, sharp objects, and potentially harmful fumes, often lacked documented safety protocols, such as mandatory use of personal protective equipment (PPE), emergency procedures, or proper ventilation. This informal approach presented potential risks to both patients and staff, increasing the likelihood of cross-contamination or workplace accidents due to inconsistent practices.

#### Patient Review and Follow-up

The continuum of care extending beyond device delivery was also largely unstructured. Follow-up protocols were informal or absent. Most facilities (n=9) lacked structured documentation guiding post-fitting review schedules or outcome assessments. Patients were often simply told to return "if they had a problem" rather than being scheduled for routine check-ups at specific intervals (e.g., 1 month, 3 months, 6 months post-fitting). The methods for assessing device functionality, patient satisfaction, or long-term outcomes were not standardized, making it difficult to systematically evaluate the effectiveness of interventions or identify areas for improvement in device design or service delivery. This informal approach to follow-up meant that potential issues with the device (e.g., fit changes, wear and tear, alignment shifts) might go undetected until they became severe, impacting patient comfort and device longevity.

##### Implications of SOP Absence

The widespread absence of SOPs carries significant and detrimental implications for the entire P&O ecosystem in Ghana. The most immediate and palpable consequence is inconsistent service delivery. Without standardized procedures, similar clinical conditions and patient needs may be managed differently across facilities or even within the same facility by different practitioners. This leads to a "postcode lottery" effect, where the quality and type of P&O care a patient receives can depend more on the specific clinic or clinician, they encounter than on their actual clinical need or an evidence-based approach. Such variability undermines public trust in the P&O sector and can lead to inequitable access to quality care.

The lack of SOPs severely limits the ability to implement effective quality control mechanisms. Without documented benchmarks for each stage of the P&O process, it becomes challenging to conduct internal audits, monitor performance indicators, or ensure comprehensive quality assurance. There are no clear criteria against which to measure the quality of a cast, the precision of a socket, or the accuracy of an alignment. This absence of measurable standards makes it difficult to identify areas for improvement, address recurring errors, or systematically enhance the overall quality of devices and services. It also hinders accountability, as there are no agreed-upon standards against which practitioner performance can be objectively evaluated.

For facilities involved in student training, the absence of SOPs creates significant barriers to effective training and knowledge transfer. Without a standardized curriculum of practical skills and documented procedures, it becomes difficult to ensure that all students receive consistent instruction and achieve uniform learning outcomes. Supervisors may teach based on their personal methods, leading to variations in student competency. This lack of standardization weakens the ability to objectively assess student progress and ensures that graduates enter the workforce with a fragmented understanding of best practices. Furthermore, the absence of written protocols makes it challenging to transfer institutional knowledge, especially when experienced staff retire or move on, leading to a continuous reinvention of processes rather than building upon established best practices.

## Discussion

The pervasive issues of equipment downtime and the absence of standardized operating procedures (SOPs) observed in Ghanaian Prosthetics and Orthotics (P&O) clinics are not isolated incidents but rather symptomatic of deeper, systemic challenges that often characterize healthcare infrastructure in low-income settings. Reflecting on the field observations, several interconnected root causes emerge, collectively undermining the capacity of these clinics to deliver consistent, high-quality P&O services.

A primary root cause is limited government funding allocated to the P&O sector. In many developing nations, including Ghana, healthcare budgets are often stretched thin, prioritizing acute care and communicable diseases over specialized rehabilitative services like P&O [14,15]. This budgetary constraint directly impacts the ability to procure new, robust equipment, invest in regular preventive maintenance, or even purchase essential spare parts [16]. When funds are scarce, the immediate operational needs often overshadow long-term sustainability planning, leading to a reactive rather than proactive approach to equipment management [17]. This financial neglect creates a vicious cycle where equipment degrades, services are compromised, and the overall P&O infrastructure remains underdeveloped.

Compounding the funding issue is the severe lack of dedicated technical maintenance teams and specialized biomedical engineers within the country [18]. The sophisticated nature of P&O equipment requires specific expertise for both routine maintenance and complex repairs [19]. As observed, the absence of locally available, trained technicians means that clinics are often reliant on external, often foreign, expertise, which is both costly and time-consuming, if available at all [20,21]. This forces facilities to operate with broken machinery for extended periods, as the logistical and financial hurdles of repair become insurmountable. The problem is exacerbated by a general shortage of training programs that equip individuals with the necessary skills to maintain this specialized equipment, creating a critical human resource gap [22,23].

Furthermore, the absence of a centralized policy framework and national protocols for P&O services in Ghana is a significant contributor to the observed inconsistencies. Unlike more established healthcare disciplines, P&O in Ghana appears to lack a cohesive national strategy that dictates clinical standards for equipment, clinical practice and quality control [24–26].

This void leads to a fragmented landscape where each clinic operates independently, developing its own ad-hoc methods based on the individual experiences of its practitioners. The World Health Organization (WHO) and the International Society for Prosthetics and Orthotics (ISPO) have developed comprehensive standards for P&O services, covering policy, products, personnel, and provision [11,12]. However, as evidenced by studies in other low-income countries like Namibia, adherence to these global standards often remains partial, particularly concerning policy and governance [27]. The lack of governmental direction and a national P&O committee in many low-income contexts directly contributes to the inconsistent and protracted decision-making that impedes service delivery [28,29].

Finally, low data reporting and inadequate inventory management further obscure the true extent of the problem and hinder effective planning. Without systematic data on equipment functionality, maintenance history, and service needs, it becomes impossible for ministries of health or facility administrators to make informed decisions about resource allocation, procurement, or training priorities [30,31]. This lack of granular data perpetuates a cycle of reactive crisis management rather than strategic, evidence-based development [32].

The challenges observed in Ghana are not unique; they resonate deeply with the experiences of other low-income countries globally. Research on assistive technology provision in regions like Sierra Leone and Malawi consistently highlights similar barriers, including a high percentage of assistive devices needing repair, difficulties in accessing repair services due to cost and distance, and a general lack of resources for sustained maintenance and follow-up care [33,34]. Studies across sub-Saharan Africa frequently report that a significant portion, up to 40% of medical equipment, including P&O machinery, is non-functional due to inadequate maintenance, lack of spare parts, and insufficient trained personnel [35,36]. This widespread issue underscores a systemic vulnerability in healthcare systems reliant on imported technology without corresponding investment in local maintenance capacity and robust policy frameworks [37].

The cumulative impact of these issues on long-term patient care is profound. Patients face prolonged waiting times, receive devices of inconsistent quality, and may experience suboptimal functional outcomes [38,39]. This not only impedes their rehabilitation and integration into society but also places an increased burden on their families and the healthcare system [40]. The cost implications are also significant; frequent repairs of poorly maintained equipment, the need for outsourcing, or the premature replacement of devices due to irreparable damage represent substantial financial drains that could be mitigated through preventive measures [41,42]. Moreover, the hidden costs of patient dissatisfaction, potential complications from ill-fitting devices, and reduced productivity due to prolonged disability are immeasurable [41–43].

The observed conditions also have a tangible effect on staff morale. P&O professionals are dedicated to improving lives, but constantly working with broken tools and without clear operational guidelines can lead to frustration, burnout, and a sense of futility [19,44]. This can hinder professional development and even lead to a brain drain, as skilled individuals seek opportunities in environments where they can practice their craft effectively [45–47]. Ultimately, patient satisfaction suffers when services are delayed, device quality is inconsistent, and follow-up care is compromised [34,48,49]. Patients lose trust in the system, potentially leading to abandonment of devices and a return to a state of reduced mobility and independence [50].

Despite the daunting nature of these challenges, the field observations underscore a critical pathway forward: standardization and basic maintenance planning can go a long way to improve the P&O field in Ghana. Implementing national SOPs for all stages of P&O service delivery in Ghana, from assessment and casting to fabrication, fitting, and follow-up, would ensure a baseline level of quality and consistency across clinics. These SOPs would provide clear guidelines, reduce reliance on individual discretion, and facilitate better training [1,51]. Simultaneously, a focus on basic, preventive maintenance planning, coupled with investment in training local technicians, could significantly extend the lifespan of existing equipment [52,53]. Even simple measures, such as regular cleaning, lubrication, and minor adjustments, can prevent major breakdowns[54]. Prioritizing the establishment of a robust supply chain for essential spare parts and fostering local repair capabilities would further enhance the operational resilience of P&O clinics [55–57]. These foundational steps, while seemingly modest, represent a powerful leverage point for transforming P&O services in Ghana, moving towards a more sustainable, efficient, and patient-centered model of care.

## Study Limitation

Although this report provides insights from all 14 officially registered prosthetics and orthotics facilities in Ghana, certain limitations must be acknowledged. Facilities operating informally or without official registration were not included, and therefore the observations may not reflect the practices or challenges within those settings. In addition, the report is based primarily on narrative field observations rather than systematic quantitative measurements, which introduces the possibility of observer subjectivity. While this approach offers rich contextual understanding, it may not fully capture the frequency or magnitude of equipment downtime across facilities. Furthermore, the focus was limited to facility-level practices, without incorporating the perspectives of patients, policymakers, or equipment suppliers, whose views could have added further depth to the analysis. Finally, as a narrative report, the findings are descriptive in nature and do not establish causal relationships, but rather highlight trends and gaps that merit further systematic investigation.

## Conclusion

The field observations in Ghanaian Prosthetics and Orthotics clinics have unveiled critical systemic gaps, most notably the pervasive challenge of equipment downtime and the striking absence of standardized operating procedures. These issues, rooted in limited government funding, a dearth of specialized maintenance expertise, a lack of centralized policy, and insufficient data reporting, collectively impede the effective delivery of essential P&O services. As demonstrated by similar contexts in other low-income countries, these are not isolated problems but rather widespread barriers to quality healthcare provision.

Addressing these fundamental systemic deficiencies is not merely an operational necessity but a moral imperative. By prioritizing the development of national guidelines and minimum SOPs, establishing robust maintenance tracking systems, investing in local technical training, actively engaging the Ministry of Health for support, and fostering inter-facility knowledge sharing, Ghana can embark on a transformative journey for its P&O sector. Solving these deeply entrenched issues will directly translate into improved care delivery, ensuring that patients receive timely, high-quality, and safe prosthetic and orthotic devices. Furthermore, it will significantly enhance training outcomes for aspiring P&O professionals, equipping them with the practical skills and standardized knowledge necessary to excel. Ultimately, these concerted efforts will bolster the long-term sustainability and resilience of P&O services across Ghana, paving the way for a future where mobility and independence are accessible to all who need them.

## Declarations

### CRediT authorship contribution statement

**Daniel Opoku-Gyamfi**: Writing – review & editing, Writing – original draft, Methodology, Formal analysis, Data curation, Conceptualization. **Akouetevi Aduayom-Ahego**: Writing – review & editing, Writing – original draft, Supervision, Methodology, Conceptualization. **Christian Kwaku Agbewoavi**: Writing – review & editing, Data curation, Formal analysis. **Cecil Owusu Bempah**: Writing – review & editing, Methodology, Formal analysis, Data curation. **Joseph Pitoor Bikala**: Writing – review & editing, Methodology, Formal analysis, Data curation. **Deborah Adiki Aniteye**: Writing – review & editing, Methodology, Formal analysis, Data curation. **Prince Oduro**: Writing – review & editing, Data curation, Methodology, Formal analysis. **Harriet Amakye Ansah**: Writing – review & editing, Data curation, Methodology, Formal analysis. **Michael Kuaku Bansah:** Writing – review & editing, Data curation, Methodology, Formal analysis. **Zenith Yamoah**: Writing – review & editing, Methodology, Formal analysis.

### Ethics statement

Ethical approval for this study was obtained from the University of Health and Allied Sciences Research Ethics Committee (UHAS-REC), under the assigned protocol ID UHAS-REC.123.33. Prior to commencing any site visits or data collection, formal permission was secured from the heads of each of the fourteen participating prosthetics and orthotics (P&O) facilities.

## Acknowledgments

We extend our deepest gratitude to the heads and practitioners of all fourteen prosthetics and orthotics facilities across Ghana who graciously welcomed our research team and provided an exceptionally conducive environment for our field observations.

## Data availability

The authors do not have permission to share data.

